# Consultagene: Pre- and Post-Pandemic Experience with a Web-based Platform and Remote Delivery of Genetic Services

**DOI:** 10.1101/2021.01.20.21249267

**Authors:** Kim C. Worley, Daniel Lee Riconda, Amy M. Wallis, Chad A. Shaw, Chris M. Holder, Salma A. Nassef, Sandra Darilek, Seema R. Lalani, Pilar L. Magoulas, Sarah M. Huguenard, Brendan H. L. Lee

## Abstract

Changes in genetics and genomics sequencing in recent years have created increased demand for genetics professionals, including clinical geneticists and genetic counselors. A significant workforce shortage of these professionals has become widely recognized. This shortage is driven by several factors, including the increased role of genetics in healthcare due to precision medicine initiatives and demand outside of medical practices in clinical and direct-to-consumer genetic testing companies that require genetics professionals for education and counseling.

We developed the Consultagene virtual platform for delivery of genetic care, counseling, and education to address some of these issues that have become global challenges in the field of clinical genetics and to bridge existing gaps at the point of care. The platform provides access to specific content based upon the referral indication including educational videos and resource links, allows document sharing, health and history information gathering and appointment scheduling with persistent access to the materials via secure data infrastructure. Having the platform coupled with access to our tele- and video consultation service allows clients convenient on-demand access to genetic education and services as needed.

This report describes the Consultagene platform development and use prior to and during the COVID-19 pandemic, some of the identified strengths and weaknesses of such a platform, and the current applications in the new environment where telemedicine practice has rapidly expanded.

**Topic Summary:** Consultagene is the first academic virtual platform to integrate a comprehensive range of genetics services including genetics education, genetics consultation, and genetic counseling. Consultagene addresses the increasing demands and unmet, evolving needs for genetic services due to widely available genomic sequencing and the platform seamlessly adapted to the pandemic-induced adjustments to clinical practice.

## Introduction

Rapidly increasing volumes of data are impacting medical genetics, where the decrease in cost and increase in throughput of sequence-based assays has driven significant change. In genetics, this clash occurs at the point-of-care and manifests in a mismatch between the demand for genetic services and the locally available capacity (Hoskovec et al., 2018). Traditional systems provide patient and provider education, patient phenotyping, genetic diagnostic evaluation, and genetic counseling via in-person, one-on-one interactions, and the required genetics expertise for the complete spectrum of care is often only available in academic medical centers in large cities.

Consultagene was conceptualized to address this mismatch by implementing a web-based platform to provide access to Baylor College of Medicine (BCM) genetics providers (physicians, genetic counselors, trainees), to expand access to BCM genetics services beyond the local Texas Medical Center clinics and to improve efficiencies within existing clinical services (see Table 1). With this platform, tele- and video-conferencing are provided for both genetic counseling and peer-to-peer consultations. The increased volume of available genetic sequence data and functional annotation also manifests in increased time and decreased workflow efficiency related to increasing education and data gathering needs. Two features of the Consultagene platform directly address these issues. First, the remote and asynchronous access to curated educational materials aims to improve workflow and allows patients to access the appropriate information in a time and place where they can focus on the material, to view the information at home with their family members and to have continual access to this information allowing them to review this information as often as needed. Second, collecting genetics-focused health history and developing pedigrees in advance of genetics visits can also improve workflow efficiency when patients can complete this information at home in advance of an appointment. The Consultagene platform was also designed to improve the access to genetics services beyond the office visit by providing continuous engagement before, during and after a clinic visit. A final motivation was to develop a solution that could provide these benefits across the continuum of care served by BCM geneticists and genetic counselors, including preconception, prenatal, pediatric and adult specialty clinics. These clinics have different workflows that require a highly flexible implementation to ensure that the needs for each clinic can be met.

**Table 1.**
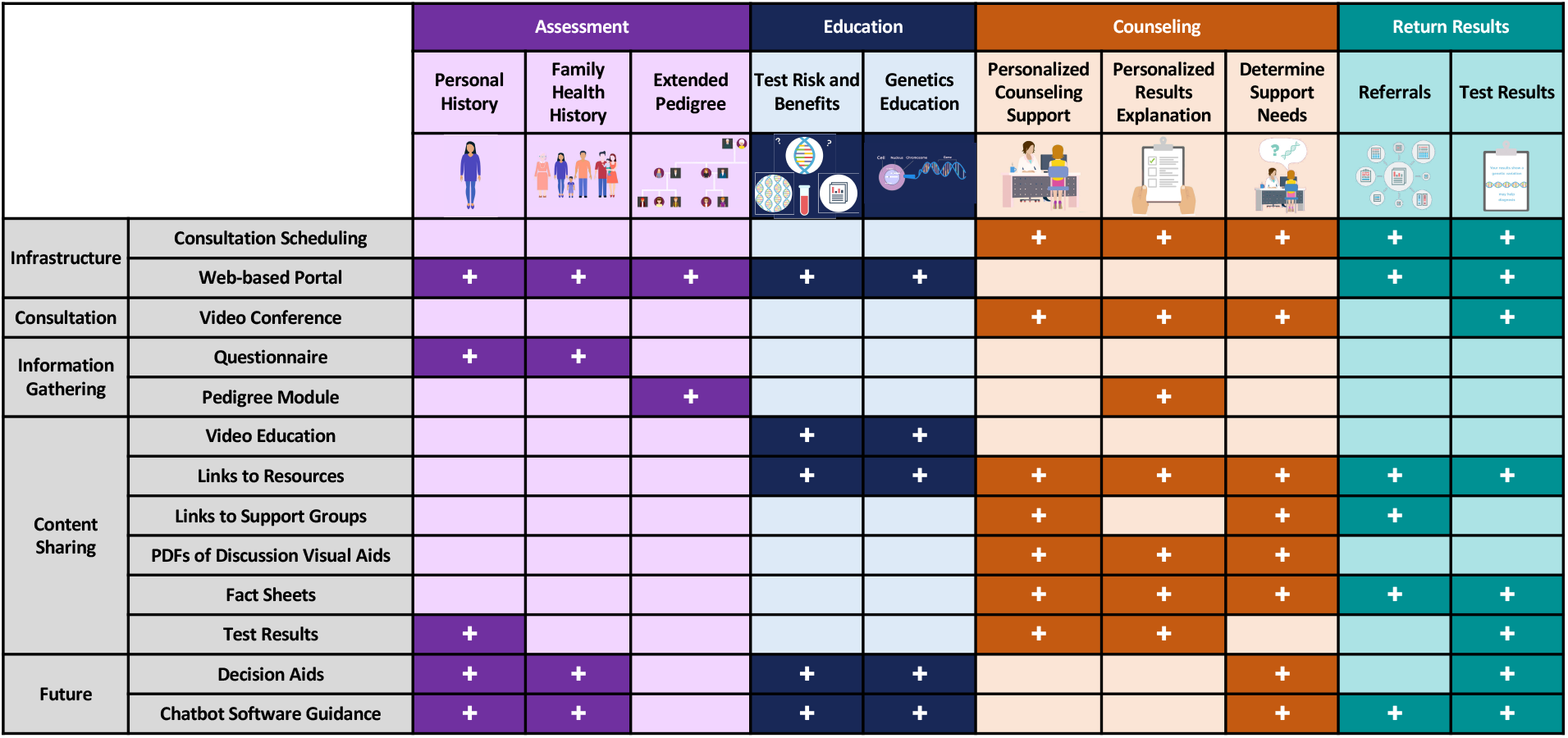
Genetic Counseling Activities Supported by the Consultagene Infrastructure.

|  |  | Assessment |  |  | Education |  | Counseling |  |  | Return Results |  |
| --- | --- | --- | --- | --- | --- | --- | --- | --- | --- | --- | --- |
|  |  | Personal History | Family Health History | Extended Pedigree | Test Risk and Benefits | Genetics Education | Personalized Counseling Support | Personalized Results Explanation | Determine Support Needs | Referrals | Test Results |
| Infrastructure | Consultation Scheduling |  |  |  |  |  | + | + | + | + | + |
|  | Web-based Portal | + | + | + | + | + |  |  |  | + | + |
| Consultation | Video Conference |  |  |  |  |  | + | + | + |  | + |
| Information Gathering | Questionnaire | + | + |  |  |  |  |  |  |  |  |
|  | Pedigree Module |  |  | + |  |  |  | + |  |  |  |
| Content Sharing | Video Education |  |  |  | + | + |  |  |  |  |  |
|  | Links to Resources |  |  |  | + | + | + | + | + | + | + |
|  | Links to Support Groups |  |  |  |  |  | + |  | + | + |  |
|  | PDFs of Discussion Visual Aids |  |  |  |  |  | + | + | + |  |  |
|  | Fact Sheets |  |  |  |  |  | + | + | + | + | + |
|  | Test Results | + |  |  |  |  | + | + |  |  | + |
| Future | Decision Aids | + | + |  | + | + |  |  | + |  | + |
|  | Chatbot Software Guidance | + | + |  | + | + |  |  | + | + | + |

## Background

The large and diverse clinical faculty of the Department of Molecular and Human Genetics (DMHG) at BCM consists of 30 practicing medical geneticists and 45 genetic counselors who specialize in a variety of settings, including: adult genetics, pediatric genetics, cancer genetics, prenatal genetics, laboratory, research, and education. In addition, there are 19 ABMGG certified molecular genetic laboratory directors who are authorized to sign out genetic testing results. The diagnostic laboratory, Baylor Genetics, produces an ever-increasing number of test results and serves clients far beyond the Texas Medical Center. Despite the large scale of the clinical activities, the types of activities required in the clinical mission at BCM and supported by the Consultagene platform are found at other genetics practices, regardless of the size.

Education and counseling about genetic testing in a clinic setting have historically been provided by genetics specialists or other health care providers during face-to-face encounters. While this is effective, it is not easily scalable and may not be the most efficient way to deliver genetic counseling and other genetics related services in an increasingly busy and time-constrained clinic environment (Barry, 2002; Holmes-Rovner & Wills, 2002). In addition, clinical specialists with limited genetics training may order genetic tests for their patients without the assistance of genetic specialists. Thus, there is a greater need to educate an increasing number of patients and providers about the rationale for genetic tests (Albada, Ausems, Otten, Bensing, & van Dulmen, 2011; Cragun et al., 2020). To bridge the gap at point of care, alternative service delivery models that engage patients and providers in unique ways are necessary to increase provider efficiency, save time, increase access to care, and provide better patient experiences (Albada et al., 2011; Buchanan et al., 2015; Castellani et al., 2011; Green et al., 2005; Griffith, Sorenson, Bowling, & Jennings-Grant, 2005; Schwartz et al., 2009; Wang, Gonzalez, Milliron, Strecher, & Merajver, 2005; Yee et al., 2014). Therefore, we developed a virtual platform for delivery of genetic care, counseling, and education to address some of these issues that have become global challenges in the field of clinical genetics and to bridge these existing gaps.

In 2015, the DMHG faculty and staff began development of Consultagene™, an online, multimedia genetics service delivery platform with the intent of improving efficiency, providing patient educational support, and delivering a variety of telehealth services (Riconda et al., 2019). The modular online platform consists of a three-party interaction among the “client”, the “referrer”, and the “genetics provider”. A referring indication specifies client “journeys” that are customized for a guided individual experience by provisioning modular components to each “journey” (see Tables 1 and 2, Figure 1).

**Table 2.**
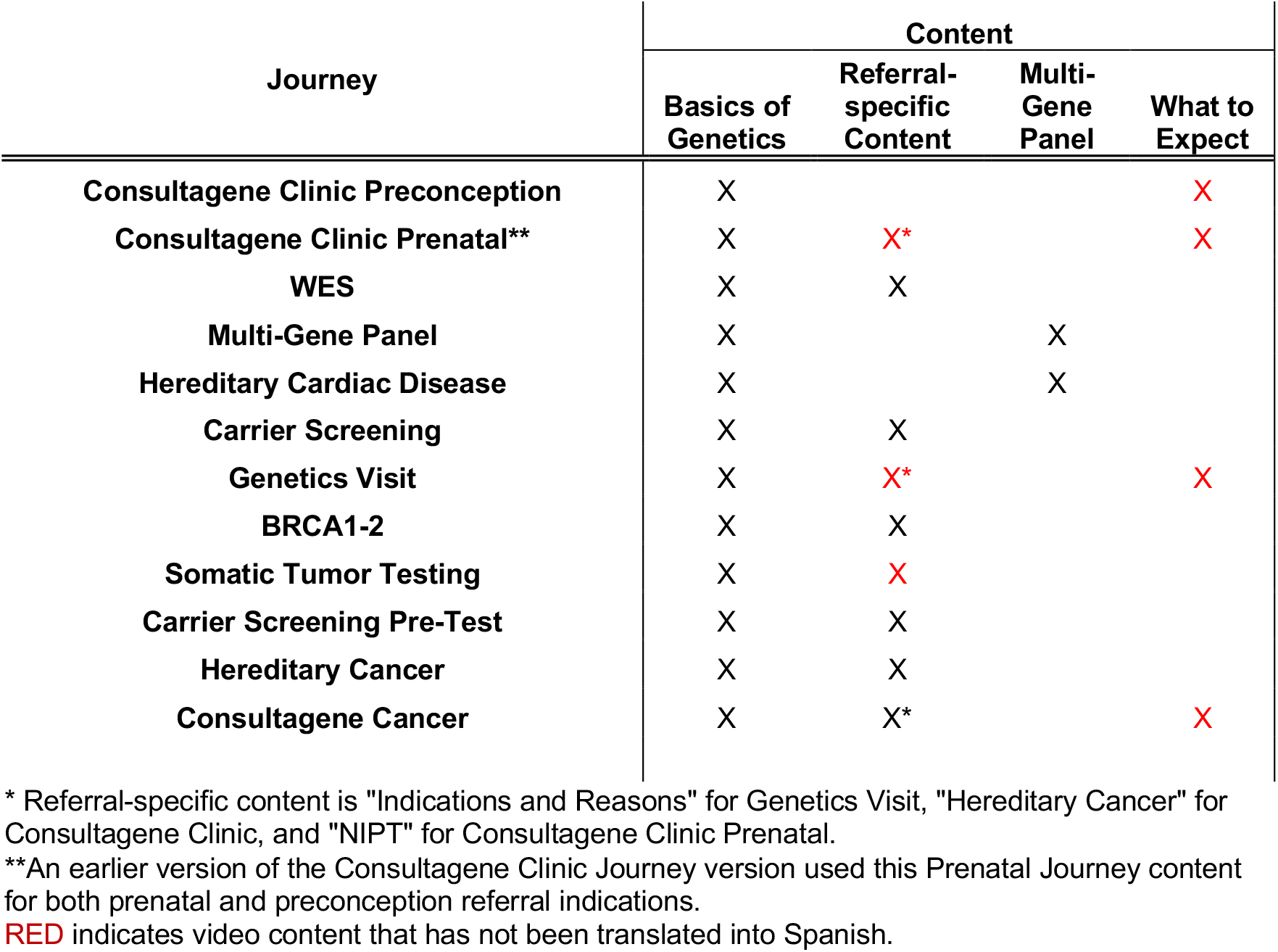
Journey Video Content Packages.

**Figure 1.**
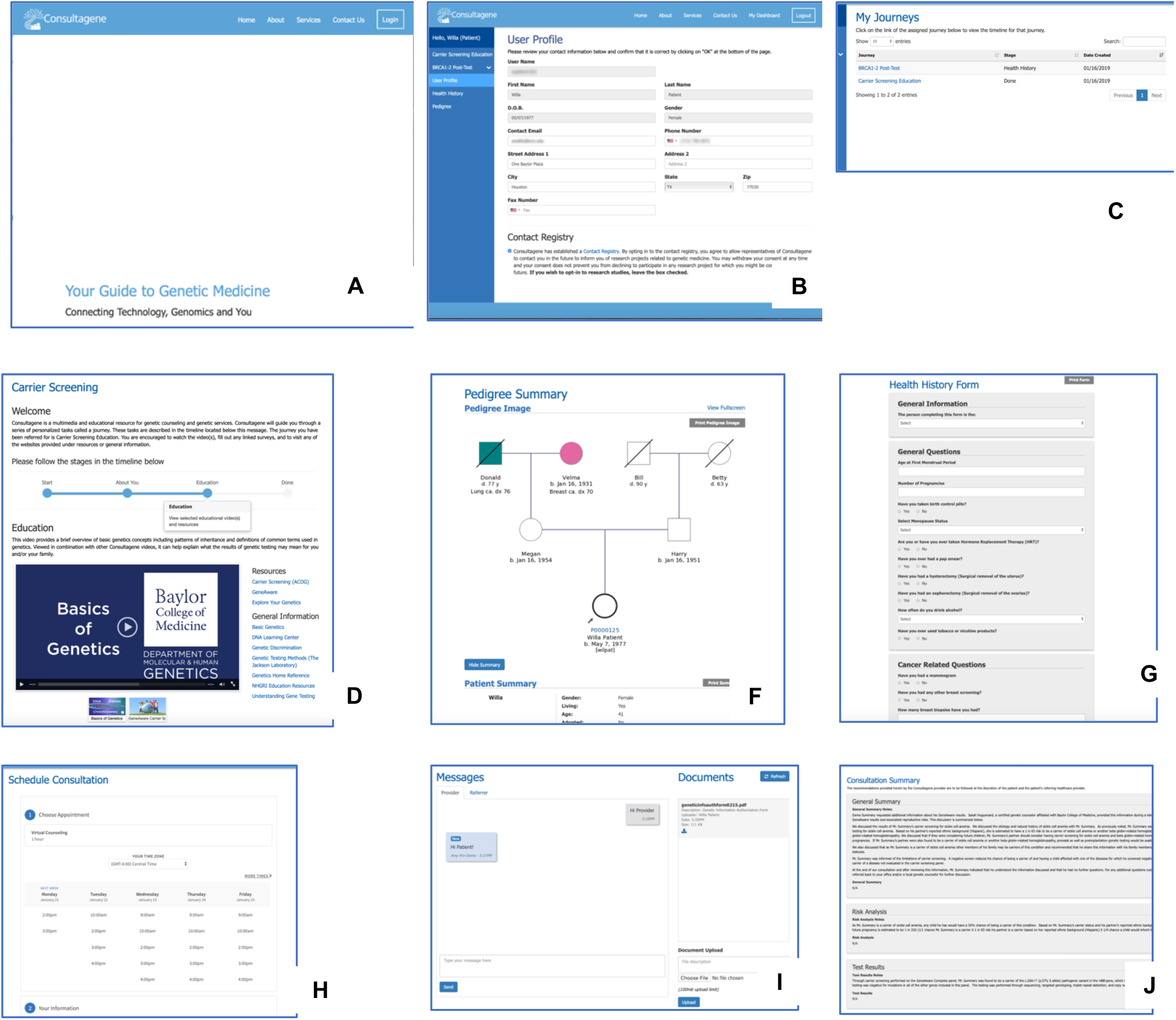
Client-Participant Consultagene Journeys. Each Consultagene Journey brings together the modules to meet the specific needs associated with the referral indication. Panels **A to F** show the **Education Only Carrier Screening Journey** for a hypothetical client. **(A)** Consultagene landing page (image of actors as clients not shown); **(B)** Login page; **(C)** Client Profile; **(D)** My Journeys Dashboard; **(E)** Journey timeline and video education screen for the Carrier Screening Education Journey; **(F)** Timeline shows the completion of the Journey. Panels **G to N** show additional modules that are a part of the **BRCA1/2 Counseling Journey. (G)** Pedigree form review screen for Client; **(H)** pedigree image generated for Client; **(I)** health history form; **(J)** post-test survey for video education; **(K)** schedule consultation; **(L)** document sharing and messaging; **(M)** consultation summary; **(N)** post-test counseling Journey completion

In February and March of 2020, the COVID-19 pandemic forced many clinical genetics centers to modify their service delivery models. The pandemic has accelerated the integration of the Consultagene platform in our prenatal, adult neurology and cancer genetic counseling service lines and modified how the platform is used in other contexts including the pediatrics practice. We describe here our experiences and lessons learned through this process.

## Methods

### Conceptual User Roles

The Consultagene experience is built around three distinct roles: the Referrer, the Client, and the Provider. The **Referrer** is the party that is requesting genetic education or counseling for themselves or a client. The **Client** is the party receiving the education and/or counseling services. The client can be a patient or a provider seeking education and/or consultation. The **Provider** is the BCM certified genetic counselor, medical laboratory director, or medical geneticist who is providing consultation to the Client either electronically or with an in-person session.

The referrer can be an individual physician who suspects a genetic component in a patient’s clinical presentation or a designated administrative representative from a physician practice that makes multiple referrals. Referrers can use Consultagene to communicate directly with both their patient and the assigned genetic counselor, and depending on the Journey, can also upload and download documents and view visit summaries. Using Peer-to-Peer Journeys (see below), physicians in the role of a client can access other genetics professionals including Medical Geneticists and Laboratory Directors

### Types of Client Interactions or Journeys

The educational and consultative path through the system is termed a **Journey**. A Journey can be a Client Journey involving the end-user client patient or a Peer-to-Peer Journey involving the referring party themselves or third-party client health provider. BCM DMHG recognized early that genetic education and counseling is necessary and beneficial to experienced care providers as well as patients and the Consultagene platform supports both.

There are three categories of Journeys: *Education Only, Consultative*, and *Peer-to-Peer. Education Only* Journeys, as the name implies are those that deliver video and other educational materials to the client without the consultation component of the Consultagene service. *Consultative* Journeys are those that, in addition to the educational materials, also include a live consultation with a genetic counselor. These can be in-person or over the integrated Zoom.us teleconferencing application. Finally, the *Peer-to-Peer* Journeys are special Journeys that incorporate the educational components and the genetic counseling, but are targeted to other healthcare professionals, including primary care physicians, specialists, or even testing laboratories. The accommodation of peer-to-peer applications allows for consultation on clinical and diagnostic topics without implicating the requirements of medical licensure.

Once a patient or physician client is referred to the Consultagene portal, the referral indication provisions the patient “Journey”, a customized guided client experience that integrates modular components. Each Journey can have one or a combination of the following components: education via video which includes pre- and post-video surveys to capture familiarity of a specific test, content, and/or effectiveness of the video as an educational tool, health history questionnaire, family history questionnaire with pedigree generation, pre-test tele-genetic counseling, post-test tele-genetic counseling, appointment scheduling, online results/records submission, and communication portal between patient, provider, and referrer. Referrers and providers can both add-on client Journeys to meet evolving needs in a client’s longitudinal care.

### Description of Client/Patient Journey Steps

As the client, patients are guided through a virtual web-based Journey which can include video education on various topics that can be tailored to the specific referral indication, such as: “*Indications for a Genetics Visit and Reasons for a Genetics Clinic Visit”, “What to Expect at a Genetics Clinic Visit”, “Hereditary Cancer”, “NIPT” and “Basics of Genetics*” (see Figure 1). For consultative Journeys, patients are asked to complete a medical health history questionnaire that is also tailored to referral indication and patient population (i.e. adult referred for cancer genetic counseling versus a child referred for a genetics clinic evaluation). Once completed, patients are further directed to complete a family history form that generates a 3 or 4-generation pedigree that can be edited and modified with family health history information. After the family history form is completed and a pedigree is generated, an email is automatically sent to the provider who will be evaluating the patient to inform them that the pedigree is ready for review. Depending on the specific Journey, the patient may also have the ability to schedule their genetic counseling session with the genetics provider through the portal, as well as upload records and any additional documentation necessary for the appointment to the provider through the document sharing section on the portal. Similarly, through the same channels, they are able to receive clinic notes and documentation from the provider before or after their genetics encounter. Each document sharing action triggers notification emails to participants. Once the patient’s Journey is completed, they have continued login access to the portal to review the videos, educational content, and resources at any time, but they are not surveyed again when they return to the portal.

### Educational Content

The BCM DMHG has developed the Consultagene platform to provide education and genetic counseling to a broad community of participants and healthcare providers. Capitalizing on the ever-growing computational resources to address limited human resources, Consultagene codifies the extensive wealth of BCM DMHG’s experience into a viable medium that is easily understood regardless of the background of the viewer. The first video, on whole exome sequencing, was scripted by genetic counselors and geneticists within the BCM DMHG and produced in collaboration with a contracted video production company. All videos have been developed and reviewed by content experts and educational specialists. Prior to release, the videos have been evaluated by naïve model users as well as genetics professional providers. Content has been improved in response to user survey feedback and to address changing practice needs such as new specialization areas (neurology) and new genetic tests (non-invasive prenatal testing). There are currently twelve Journeys (see Table 2). A typical Journey combines the “*Basics of Genetics*” video with referral specific content and the “*What to Expect at a Genetics Visit*” video if there is a consultation included in the Journey.

BCM genetics professionals engage physicians and patients from around the world and also serve a number of local patients whose native language is not English. We routinely offer counseling in both English and Spanish and because of the nature of our diverse patient population, we have become facile in using translation lines when needed for counseling in different languages. Most of the educational videos used in Journeys listed in Table 2 have been translated into Spanish. “*Basics of Genetics*” and one or two others are also available in Japanese and Mandarin respectively (Supplemental Table 1).

### Platform Infrastructure

The Consultagene platform was built using scalable enterprise computing servers running the RedHat Linux operating system. The entire infrastructure resides on a VMWare powered virtualization platform to scale as utilization demands, including to a cloud-based platform if needed. On this infrastructure foundation, the IBM WebSphere Application Server provides the secure, enterprise-class application server framework and the DB2 Database Server, and Security Directory Server (SDS) components seamlessly integrate to form a reliable, scalable, and secure environment to host the Consultagene application. We have created a secure LDAP platform for Authentication, Authorization, and Accounting (AAA) using the SDS. As a standalone product, Consultagene pulls user and group membership data from our dedicated SDS instance to govern access and authorization to only the specific areas of Consultagene needed to facilitate the user role. The WebSphere software supports the use of Security Assertion Markup Language and OAuth to federate with external providers for a Single Sign-On relationship. The secure IBM DB2 Database platform that provides the underlying data management for Consultagene supports encryption of data elements both in transit and at rest.

The IBM WebSphere Application Server is the web front end and provides User Interface and Application Programming Interface (API) access to Consultagene. These access methods are partitioned by role and further by the Referrer-Client-Provider relationships. Each function and API call made by the Consultagene codebase is made within the context of the logged-in user and is vetted against the AAA mechanisms of the SDS server, which also provides an auditing mechanism so that access and use of the system are tracked at a user level.

Through the back-end API system, Consultagene integrates with several on-premise and external Software as a Service providers, including Zoom.us (for tele- and video-conferencing), SurveyMonkey (for surveys), and Acuity Scheduling. Each of these integrations takes place through Secure Socket Layer encrypted channels for the alerting and data acquisition mechanisms of the integration.

Acuity Scheduling provides self-service appointment scheduling for Consultagene clients. Genetic counselors in our network maintain their available office hours in Acuity and have the option of integrating with their Office 365 Calendars as well. This provides the most up-to-date availability for clients needing to schedule consultation appointments. Once the client is alerted that they have progressed to a point in their Consultagene Journey where they are ready to meet with the Genetic Counselor, they simply click on the Scheduling link in their Journey and the Acuity self-service scheduling page is presented to them through the Consultagene user interface. When they have chosen their desired appointment time, they receive a confirmation email from Consultagene with their appointment information. For virtual appointments, this includes an automatically-generated, unique Zoom.us teleconference link that is created through the integration between Acuity and Zoom.us. On the Provider side, the Genetic Counselor also receives a notification that an appointment has been scheduled and if they are using the Acuity-Office 365 integration, the appointment will be automatically added to their calendar.

### Platform processes

The Consultagene platform interaction begins with a referral. Consultagene can receive referrals through our secure web-based application (Figure 1.A) or via an automated application programming interface (API) from an authorized institution. The details in the referral are used to provision a client account (for a new client – either the referred patient or provider receiving Consultagene services) and instantiate the requested Journey for that client (Figure 1.B, 1.C). As a client’s needs evolve, subsequent Journeys can be added to a Client’s profile as necessary to address the needs.

For new clients, an encrypted URL is generated with an embedded security token and automatically sent to the email address the client provided at registration. When launched, this secure token is validated by the Consultagene application and the user is prompted to validate their identity with a personal vital statistic provided directly from their intake form. Once validated an automatically generated username is assigned to the user and they are prompted to create their password.

When an existing patient is being assigned an additional Journey, the system automatically generates an email notification of the addition and provides a link to Consultagene. As the patient progresses through their assigned Journey, milestone notifications are sent to the relevant parties (Figure 1.D). For example, when a client reaches the point in their Journey where they are ready to schedule their live consultation, notifications are sent to the provider and the client to inform them of the readiness and directing them to the proper resource on Consultagene.

### Surveys

At three key milestones in the Journey, patients are surveyed to get an indication of the patient’s level of understanding, to gauge the effectiveness of the educational material, and to review the Consultagene experience. Surveys are initiated before patients view the educational videos, when they are asked to indicate their level of understanding of key concepts related to the Journey. After viewing all of the videos a companion survey is offered to assess the efficacy of the education. Finally, as the patient completes their Journey the final survey is emailed to them to solicit feedback on the overall Consultagene process and experience. All of these surveys are anonymous and the platform does not capture any identifying data such as IP address or name.

### Information gathering

The health history and pedigree modules are specifically designed for information gathering (Figure 1.F, 1.G). Each provides a specific questionnaire to the patient to collect information related to the Journey. Both health history and pedigree forms have specific versions for cancer and non-cancer indications. A common health history is maintained for the patient and is available for add-on Journeys. When the Consultagene platform was initially designed, few of the affiliate sites subscribed to the Pedigree module in their commercial EMR system and the pedigree information collected by the integral module was more limited than pedigrees required for genetic counseling and genetic evaluation. For this reason, a platform specific pedigree module was developed, which continues to be used. The Consultagene pedigree questionnaires solicit information for up to four generations. As with the health history, specific cancer and non-cancer versions are used. Genetic professionals can update and edit the pedigrees directly in Consultagene through the customized view into PhenoTips provided with the application. The edited pedigree information can be pushed into the EMR as a .pdf report.

### Document sharing and messaging

Document sharing is supported by the Consultagene platform and is utilized for information gathering from the Client and results reporting or supplemental education materials from the provider. Messaging through the platform supports direct requests from any role (the Provider, Client, or Referrer). Requests for additional relevant information are executed through an email from Consultagene and response information sharing through secure messaging within the platform (see Figure 1.I).

### Summary and continued access

The final step in a consultative Journey (both for Clients and Peer-to-Peer) is the document Summary (Figure 1.J). This Summary document is completed by the Provider and encapsulates the information discussed during the consultation session. It is broken down into logical segments that cover the general areas of discussion from Test Results to Risks and Next Steps. Each individual section of the Journey Summary document contains its own narrative as well as other electronic media that can be attached to that section. After viewing and acknowledging the Summary, patients are informed that their Journey has come to its completion and are reminded that, though this Journey has ended, they will retain access to all of the materials in perpetuity.

### Experience of the Practice Referrer

Practice referrers initiate the Consultagene process, by completing a web form or submitting an API call providing information about the client and the indication for the referral. This information informs the type of journey and interaction the client will have with the platform and service. If a consultative journey is selected, the referrer receives documentation summarizing the consultation following its completion. The referred has access to a list of the patients for whom they have requested a journey, has access to the messaging function (described above) which allows them to have any additional needed communication with the provider, and can follow a patient’s progress through the journey.

### Experience of the Provider

Genetic Counseling providers within the Consultagene portal are assigned patients and can have access to a list of patients for whom they provide genetic consultations. The genetic counseling provider is able to monitor the progress a patient has made within their Journey, including completion of video education, intake paperwork, and family history and also has access all forms the patient has completed or records the patient has uploaded as part of their Journey. Once a genetic consult has been provided, the providers are able to document their consultation notes in the portal, which are then available to the patient and the referring provider. In addition, the provider has the ability to upload visual aids or patient education resources, making these readily available for patient access. The portal allows for messaging between the genetic counselor and the patient as well as between the genetic counselor and the provider. All records collected through the Consultagene platform are available for transfer to another medical record system.

## Results

Consultagene initially went online at the end of 2018. Usage has varied over time with slow growth at first, rapid growth in the middle of 2019 and a drop in use in early 2020. Between May 2019 and January 2020, ∼150 Journeys were created per month. Following the local official COVID-19 pandemic-related shut down measures beginning on March 11, 2020, the volume decreased to ∼75 Journeys per month.

Patient experience with the platform varies by Journey. Reports from surveys consistently indicate increased understanding and perceived value. The user experience survey questions from the most frequent Journeys are summarized in Figure 2, the raw survey data is in the Supplemental Excel file (CG Survey Data) with the demographic information of the survey participants in Supplemental Figure 1,. The wording of questions on the surveys for different Journeys are slightly different, but more than 330 patients responded to each of the questions shown in Figure 2 and the majority either agree (32 to 56%) or strongly agree (31 to 42%) with statements that the videos were easy to understand, provided a good explanation, were helpful and informative and that the patient’s understanding improved after watching. For exome sequencing (WES), prior to viewing the video, familiarity with WES was 1.77 on a 1 to 5 Likert scale (1 = not familiar; 5= very familiar) compared to 4.09 (having a better understanding) and 4.12 (the platform provided a good explanation of WES) after viewing the video. These data suggest that the video education in the context of a virtual integrated delivery platform is an effective tool that increases knowledge and understanding.

**Figure 2.**
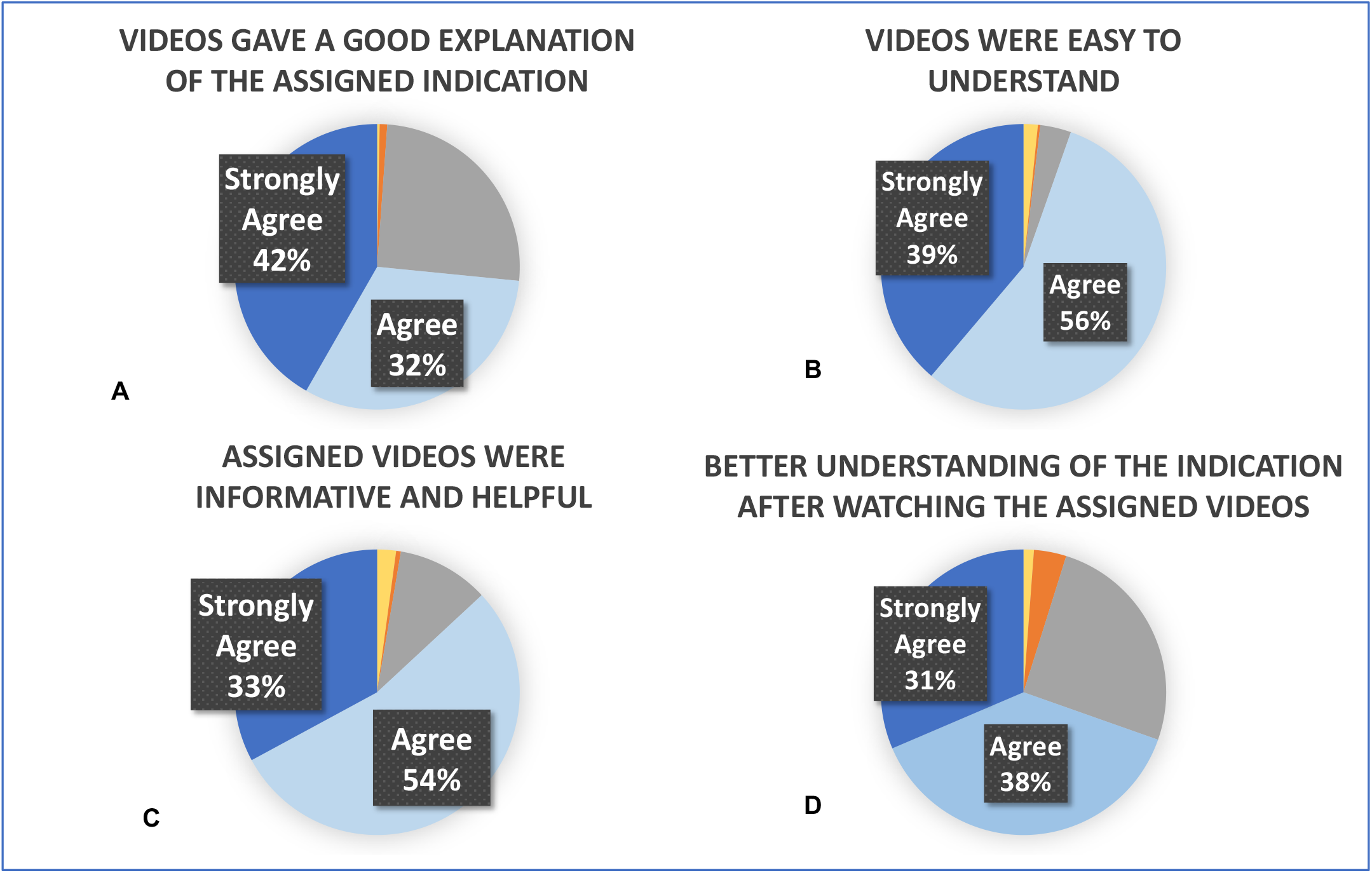
Experience Surveys. Surveys of client experience are shown with each statement that respondents were asked to score indicated at the top. Responses were scored on a 5-point Likert scale from Strongly Disagree (yellow), Disagree (orange), Neutral (grey), Agree (light blue), to Strongly Agree (dark blue). In all cases, the majority of the survey respondents found the material understandable and informative. (**A**) Total responses 411, the majority from the WES Journey (367), with the remainder from cancer surveys (44). Overall, 61 (15%) opted out. (**B**) and (**C**) were on the same survey with total responses 375, the majority from the Basics of Genetics Journey (335), with the remainder from hereditary cancer (29) and NIPT (11). Overall, 1 (0.3%) opted out. (**D**) Total responses 742, including 367 from the WES Journey, 331 from the Basics of Genetics Journey, and the remainder from cancers (43) and NIPT (11). Overall, 61 (8%) opted out.

Consulting clinical geneticists also find that the platform saves time and increases efficiency. In a test cohort of patients (n=30) for a pre-specified “Genetics Visit” Journey to be completed prior to an in person clinical genetics evaluation at Texas Children’s Hospital, patients were guided through a virtual web-based Journey. This Journey included video education on three topics: “Indications and Reasons for a Genetics Clinic Visit”, “What to Expect at a Genetics Clinic Visit,” and “Basics of Genetics”, completion of a medical health history, and completion of a family history form that generates a pedigree prior to the clinic visit. Once the steps were completed, an email notification was sent to the provider who was able to view the completed forms and pedigree before the clinic visit. Provider surveys (n=11) indicated that obtaining the health history and pedigree before the clinic appointment saved time, increased their efficiency during the clinic visit, and was preferable to previous workflows when all these activities occurred during the clinic visit.

The use of Journeys in Consultagene has changed over time. Initially Education Only Journeys were planned to have no-provider interaction but to rely on video and online resource education only. Pre-Test and Post-Test Journeys both included the consultation steps of provider assignment, health history, pedigree, scheduling, consultation, summary with communication. Genetics Visit included provider assignment, health history, pedigree, and with communication. With use, the concept of education only and add-on Journeys has evolved as patient needs are not linear and pre-and post-test Journeys have been specialized to be less redundant.

### Evolution of In-Person versus Tele-Genetic Counseling

The formation of the Consultagene Clinic in early 2019 resulted from the transition of a traditional in person genetic counseling service to a hybrid model allowing for both in person and telehealth consultations. The clinic leverages the Consultagene platform to provide pre-consultation education along with a mechanism for patient and provider communication and record sharing. All patients receiving a consult, regardless of mode of delivery, are given access to the platform and assigned a Journey. For all patients seen within the practice, documentation of the consultation is housed within the Consultagene portal and can be accessed by both the patient and referring provider.

Prior to COVID 19, about 50% of patients elected telehealth consults, anecdotally citing reduction of travel time and parking fees, while allowing more flexibility for patients and their family members to coordinate appointment scheduling as their primary factors for electing this modality. In response to COVID 19, the practice immediately shifted completely to telehealth, with minimal impact on patient volume.

The fact that the Consultagene platform was in use at the time that BCM stopped in-person clinical visits in mid-March 2020 due to COVID greatly facilitated the transition to tele-health. Our counselors are in the unique position of serving prenatal patients both through the Consultagene Clinic and through the Texas Children’s Hospital (TCH) Pavilion for Women and experienced differences in implementing telemedicine during this time. Prior to the COVID-19 pandemic, TCH genetic counseling visits were conducted in person, while Consultagene Clinic visits were a combination of in-person and video consultations. The Consultagene Clinic transitioned immediately to 100% tele-health, while the TCH transition was a multi-step process. The Consultagene Clinic transition to telehealth visits was seamless since there was already the availability and experience with video consultations and patient intake paperwork was available for patient completion through the Consultagene platform. The workflow of the Consultagene virtual clinic experienced only minimal changes in response to the pandemic. In contrast, the TCH prenatal genetics clinics initially converted to phone calls for counseling. This approach presented the limitations of lack of verbal cues and access to visual aids during counseling and no access to patient intake paperwork prior to the consultation. The second TCH iteration was the conversion to video counseling, with a protocol that has been revised several times since its initiation. The transition from in-person to tele-health also varied by practice. The TCH prenatal practice had already been providing tele-health visits to select remote clinic locations, so the processes were familiar to at least some practitioners. The transition there was accomplished more quickly than for the pediatric genetics practice, likely due to the lack of an established telehealth program and the complexities of completing a pediatric genetics physical and dysmorphology examination via telemedicine. The Consultagene Clinic transition to fully tele-health consultation has led to some unexpected benefits for clinic processes. Scheduling has become easier as the timing of the visit no longer had to be coupled to another doctor visit to minimize the need for the patient to make time in their schedule for multiple in-person visits. Additionally, new consult services for distant locations (Dallas, San Antonio, Austin) have been added, and although expansions like these were a part of the long-term plan, the mitigation guidelines implemented as a result of the COVID-19 pandemic have led to greater patient uptake and acceptance of remote consultations. The pandemic has also led to increased utilization of education-only Journeys, especially for hereditary cancer indications. In the months since the beginning of the pandemic, modifications to patient messaging have resulted in a greater number of patients completing of all the steps in the education Journey as well as improved completion rates for pre-visit paperwork. Some of these modifications were driven by need, as incomplete paperwork could no longer be handed to the patient arriving for an in-person visit.

## Future directions

With our Consultagene platform and clinic experience to date, we have identified several opportunities for growth and improvement. The improvements address both content and changes in the user experience.

More complete language support for non-English languages is planned. Although the educational materials and videos are provided in both Spanish and English, the platform with login, navigation and scheduling information is thus far only available in English. Implementing the platform in Spanish will improve patient engagement for Spanish speakers without written English proficiency. This change addresses a community need in our local county, where Hispanics make up 43% of the population (U.S. Census Bureau, 2017) and account for 82% of the births in the safety net Harris Health System hospitals (one of which is staffed by BCM), and where nearly half of households (44%) speak a language other than English at home (U.S. Census Bureau, 2017). In addition, providing the platform in Spanish will enable us to improve genetic services access to underserved areas of the state with a greater proportion of Spanish speakers than Houston, including regions where BCM has existing community engagement and education activities such as Brownsville and El Paso with 81% and 94% respectively Hispanic, and 86% and 69% respectively speaking a language other than English at home (U.S. Census Bureau, 2017).

Access to specialty genetics counseling services is a second area of planned growth for Consultagene. Consultagene can serve as a centralized hub for patients to obtain consultations with genetic counselors having varied expertise. Whereas genetic counselors who provide services in medical subspecialties are usually embedded in separate departments, including neurology (for provision of neurogenetic counseling) and internal medicine (for provision of cardiovascular genetic counseling), Consultagene genetic counselors with specialty expertise are, by contrast, available on a single platform. Direct access to genetic counselors avoids long wait times for genetic counseling visits in medical specialty settings like neurology and cardiology, as these clinics experience patient demand for higher priority visits that overwhelms administrative staff, and extend the time required to triage and schedule visits with a genetic counselor among visits with other clinicians. Furthermore, most community-based neurology and/or cardiology clinics do not have a genetic counselor embedded in their practice. In these clinics, patients who require genetic counseling are referred to general genetics clinics, typically situated in large medical centers, which also experience outpatient access delays due to limited clinician resources. With Consultagene, direct access to genetic counselors with specialty expertise enhances the clinical care neurologists and/or cardiologists provide to their patients by minimizing the lead time occurring between their recommendation for genetic counseling and the visit with a genetic professional.

In addition to supporting clinical genetics activities, the flexible Consultagene design can enhance the two other academic missions – Education and Research. Education is a central mission of academic centers. The DMHG archives recordings of the ABMGG training lectures, clinical genetics lectures, genetics grand rounds, and exome and genome sign-out conferences and makes these recordings available to BCM trainees to enhance the local educational experience. The DMHG also sponsors a community educational seminar series called “Evenings with Genetics”. The BCM genetics providers are engaged in these educational efforts as well as with several patient and family support organizations for rare and/or genetic diseases. The flexibility of the Consultagene platform affords the opportunity to incorporate these specialized educational resources and to more directly target healthcare providers’ continuing education and patient education to address the needs of participants and to support these communities.

Supporting genomics and genetics research was another motivating principle of Consultagene development and BCM researchers are also beginning to adopt the platform. Genetics research that includes genomic testing and return of results as well as ongoing patient engagement are research protocols that can be well served by the Consultagene infrastructure. The flexibility of the platform can support both general needs of education about research participation as well as study-specific engagement activities including education about the study and the informed consent process. The expanding integration of virtual health in US medical offices and hospitals affects clinical research, which recruits subjects from patient cohorts characterized in the clinic. As clinical visits have transitioned to a virtual platform, so too have clinical research visits shifted. Consultagene is poised to facilitate virtual clinical research visits, including meetings between subject and research coordinator to discuss study goals and procedures, as well as risks and benefits of participation. The meeting concludes with the subject’s completion of an informed consent form. Historically completed in person, aided by accompanying phone conversations and back-and-forth email communications with the research coordinator, consent activities could instead happen via the Consultagene platform. Both general and study-specific research journeys could be viewed synchronously during a virtual meeting with the research coordinator or asynchronously at the subject’s convenience. A general research Journey would be developed with educational videos pertaining to human subject research and other content about research participation and informed decision-making intended for all potential research subjects to view. The general research Journey could be paired with customized study-specific content, intended for subjects of a particular study to view. The research journey would conclude with the completion of an electronic informed consent through Consultagene. As with current education and genetic counseling Journeys, research Journeys would support the back and forth communications, provide a persistent record of the relevant information and shared documents, and allow for continued enrolled participant engagement activities such as the return of research results and counseling for genomic research findings.

We are also working on improvements that will include a seamless integration with the Electronic Medical Record (EMR) systems used by affiliate institutions. With the Consultagene capability to use APIs to pull information from the EMR and push information to the EMR, this future development is targeting a more integrated approach. This would allow the provider to pull information from the EMR to initialize the pedigree development and to push the completed and discussed pedigree back to the EMR as structured data, to allow for more integrated scheduling, and to push the consultation documentation to the EMR.

Another goal is to foster cascade engagement through the platform. Often the information provided to a genetic counseling patient is relevant to the patient’s family (e.g. enabling cascade testing for cancer risk variant). Additionally, information from a family member can inform the patient’s diagnostic process. For example, more distant family members may have undergone testing that could inform the patient’s diagnosis or modify test result interpretation if more information about the test and the diagnosis of the family member was available. Reaching out to family members on behalf of the patient via the platform and removing this burden from the patient may improve the engagement of family members, which could benefit both the patient and the family member and increase the participation in cascade testing.

Finally, a number of process improvement opportunities have also been identified. Creating a dynamic list of action items and adding each survey and each video as more granular and integral steps in the Journey may improve the survey response rate and also aid the client’s progress through the Journey. Integrating digital assistant technology is planned to assist with common questions and aid in client interactions. And incorporating API interactions with the EMR as detailed above will allow structured information to be shared as data and metadata rather than as .pdf file attachments.

## Conclusion

Based on our experience in the development and implementation of both the Consultagene platform and Consultagene clinic we believe that expansion of alternative service delivery models, particularly leveraging online platforms for virtual genetics care engagement, may help to bridge the increasing mismatch between supply and demand for genetic services both virtually and in the in person clinical visit. Alternative delivery models, such as this platform, will be necessary given the limitations in the genetics workforce and as complex testing in the genetics clinic and beyond continues to evolve.

## Data Availability

The survey data reported is provided in the supplemental information. The Consultagene clinic can be reached via the web site www.consultagene.org. Contact the authors for information about adopting the Consultagene platform.

## Author Contributions

All the authors reviewed the manuscript, KW, BL, DR, SD, and SN wrote the manuscript, DR, SD, PM, AW, SN, and SL edited the manuscript. The platform was designed by BL, DR, CH, AW, and CS and platform content was developed by DR, PM, BL, AW, SD, SH, and SN. AW and CH compiled the survey data and AW and KW produced the figures for the manuscript. Authors KW, AW, and BL confirm that they had full access to all the data in the study and take responsibility for the integrity of the data and the accuracy of the data analysis. All the authors gave final approval of this version to be published and agree to be accountable for all aspects of the work in ensuring that questions related to the accuracy or integrity of any part of the work are appropriately investigated and resolved.

## Acknowledgements

We thank Element Blue, our partner for platform development, and our primary video vendor Wire Buzz, and other video vendors Adcetera and Kyle Lamar Productions. The development of the platform was funded with departmental funds.

## Compliance with Ethical Standards

### Conflict of Interest

The Baylor Genetics Laboratory is a joint venture between the Department of Molecular and Human Genetics at Baylor College of Medicine and H.U.Group Holdings, the authors are faculty and employees in the department and benefit indirectly from this arrangement. The authors declare they have no other conflicts of interest with this work.

The study was discussed with representatives of the Baylor College of Medicine Institutional Review Board. **No informed consent was required** from subjects as data were anonymously collected from the surveymonkey system. All procedures followed were in accordance with US Federal Policy for the Protection of Human Subjects.

No **non-human animal studies** were carried out by the authors for this article.

